# Treatment Gaps Among Young Medicaid-Enrolled Children with Tooth Decay in Pediatric Primary Care

**DOI:** 10.64898/2026.07.23.26357672

**Authors:** David Selvaraj, Sarah D. Ronis, Jeffrey M. Albert, Johnie Rose, Suchitra Nelson

## Abstract

**Objective:** To examine whether Medicaid-enrolled preschoolers with untreated decayed teeth received dental treatment within one year of enrollment and identify the factors associated with a treatment gap.

**Methods:** A retrospective cohort analysis of data from a cluster-randomized trial conducted in 18 community-based pediatric primary care practices in Northeastern Ohio (2017–2022). Treatment receipt was determined using Medicaid claims, with treatment gap defined as fewer teeth with treatment claims than teeth found on baseline exam with decay. Multivariable logistic regression assessed association of treatment gap with child age, sex, race/ethnicity, caregiver education, and number and location of baseline decayed teeth.

**Results:** Of 766 eligible children, 487 (63.6%) attended the dentist within one year. Among 155/487 (31.8%) with baseline untreated decay, 90/155 (58.1%) had a treatment gap. Odontograms visually showed that decay was concentrated on upper anterior and posterior teeth. A treatment gap was associated with a greater number of decayed posterior teeth (OR = 1.90, 95% CI: 1.60–2.30) and decayed anterior teeth (OR = 2.19, 95% CI: 1.51–3.39), both p < 0.001. Other socio-demographic variables were not significantly associated with a treatment gap.

**Conclusion:** More than half of Medicaid-enrolled children attending well-child visits had a dental treatment gap after 1 year. This pattern may reflect dentist’s hesitance to restore primary teeth nearing exfoliation and needing multiple dental visits to complete needed restorative treatment. To address this gap, non-surgical interventions such as silver diamine fluoride can be applied by pediatric primary care providers to control the bacteria and prevent disease progression.

**What’s Known on This Subject:** Dental caries is the most prevalent chronic childhood illness, disproportionately affecting low-income Medicaid-enrolled children. Despite barriers to dental access most research focuses on utilization rather than on whether children with untreated decay received needed treatment once they attend a dentist.

**What This Study Adds:** Among Medicaid-enrolled preschoolers attending well-child visits, 58.1% had a dental treatment gap for untreated decay. These findings underscore the need for greater adoption of nonsurgical silver diamine fluoride treatment that can be applied by medical professionals to close the gap.

**Contribution Statement:** Dr. David Selvaraj and Dr. Suchitra Nelson conceptualized and designed the study, drafted the initial manuscript, designed the data collection instruments, coordinated and supervised data collection, collected data, carried out the initial analyses, and critically reviewed and revised the manuscript.

Dr. Sarah Ronis drafted the initial manuscript and critically reviewed and revised the manuscript.

Dr. Jeff Albert conceptualized and designed the study, coordinated and supervised data collection, carried out the initial analyses, and critically reviewed and revised the manuscript.

Dr. Johnie Rose conceptualized and designed the study and critically reviewed and revised the manuscript.

All authors approved the final manuscript as submitted and agree to be accountable for all aspects of the work.

## Introduction

Dental caries or tooth decay is the most common chronic childhood illness in the US, affecting 23% of children 2-5 years old.^1^It is a preventable disease that, if left untreated, can lead to pain, infections, and even the rare occurrence of death.^2^ Early and regular dental visits starting by age 1 and continuing every 6 months are recommended by the American Academy of Pediatrics (AAP), the American Academy of Pediatric Dentistry (AAPD), and related professional organizations.^3,4^ However, some dentists are hesitant with dental visits starting at age 1,^5^ and hence do not follow these guidelines, resulting in only 27 % of children aged 1 to 2 years and 51 % of children aged 3 to 5 years receiving an oral examination.^6^ Early dental visits provide an opportunity for timely detection and prevention using nonsurgical caries treatments such as fluoride varnish for prevention and remineralization, and Silver Diamine Fluoride (SDF) for caries arrest.^7^ Pediatric primary care providers are also well-positioned to deliver these interventions,^8^ as the majority of young children attend well-child visits even when they do not access dental care.^9^

Even when attending dental visits, many individuals with low income may not receive needed dental treatment at all.^10^ Several dentist-related factors can contribute to such treatment gaps. The lack of mandated, standardized dental diagnostic terminology^11^ and inconsistency among dentists in use of visual exams and x-rays for caries diagnosis.^12^ introduces variability in caries diagnosis. Further, outpatient dentists may be unwilling to treat young children with complex restorative needs.^13^ Finally, there are no clear guidelines from the AAPD on whether to treat decayed teeth that are likely to exfoliate soon,^14^ such as the anterior in children aged 6-8 years.^15^

Parent-related factors that contribute to a treatment gap include: parents’ lack of knowledge to take the child to the dentist,^16^ finding a Medicaid-accepting dentist,^17^ and lacking transportation.^17^ Even when parents overcome barriers to access, minority and Medicaid-enrolled children often receive lower quality dental care.^18^ In qualitative interviews, caregivers report long wait times, negative experiences with dentists and staff, and discrimination based on race and Medicaid status.^18^ Parents often must take their child to multiple appointments to get restorative treatments completed as children can only tolerate treatment in one quadrant of the mouth per visit in the outpatient setting^19^. For extensive treatment needs, i.e. decayed teeth in several quadrants, the Operating Room (OR) under general anesthesia (GA) is frequently used.^20^ These challenges may further exacerbate oral health disparities.^21^

While a recent systematic review describes factors affecting dental utilization^17^ and we have previously reported on utilization of dental services in this cohort,^22^ to our knowledge, no literature exists examining the dental treatment gap in young children in primary care or the factors affecting it using both clinical and claims data. Therefore, this study investigated, (1) whether among Medicaid-enrolled preschoolers a dental treatment gap occurred for untreated decay, and (2) whether this treatment gap was associated with factors such as the child’s race, age, sex and location of the decay.

## Methods

### Study Design

This retrospective cohort study used secondary data collected as part of a cluster Randomized Controlled Trial (cRCT). ^23^ Only a subgroup consisting of those who had continuous Medicaid enrollment for one year after baseline, completed the baseline clinical dental exam for caries (decay) in the original cRCT, and had evidence of going to the dentist from Medicaid claims were included in this study.

The original cRCT investigated the impact of multi-level interventions to increase dental care utilization among Medicaid-enrolled preschoolers. The interventions included practice-level changes to the electronic medical record (EMR) and provider-level (pediaticians, nurse practitioners) training in theory-based didactic and skills training to deliver oral health information based on the common-sense model of self-regulation,^24^ a dental prescription, and a list of Medicaid-accepting dentists to parents. All participants (parent-child dyads and providers) were consented. More details on the design of the main trial have been previously published.^23^ The study adhered to the STROBE ^25^ guidelines for reporting observational research.

### Setting

This study took place in 18 community-based pediatric primary care practices in Northeast Ohio. Data were collected from November 2017 to August 2022.

### Participants

Children were 3-6 years old at the time of recruitment, continuously enrolled in Medicaid, and were attending a Well-Child Visit (WCV). Selection criteria for parents included acting as the child’s legal guardian, age of at least 18 years, English proficiency, and residence in the study area (northeast Ohio) for at least 2 years from the baseline. Parent-child dyads received a $40 incentive as compensation for their time. Participants who withdrew were excluded from the analysis.

### Data Collection

#### Socio-demographics

Sociodemographic information was collected via a parent self-reported questionnaire completed at the time of recruitment with items adapted from the National Health and Nutrition Examination Survey (NHANES).^26^ The items included in the present study were child sex (male or female), child race/ethnicity, collapsed to non-Black non-Hispanic, Black non-Hispanic, and Hispanic, caregiver education (>High School, <= High School) and the child’s age in years.

#### Baseline Dental Exam Data

Child’s untreated decayed tooth status were evaluated using clinical dental examinations during the baseline well-child visit. Six licensed dental hygienists utilized the International Caries Detection and Assessment System (ICDAS)^27^ to visually examine teeth without magnification or radiographs. Before examination, the teeth were cleaned with a toothbrush and dried with gauze. The ICDAS codes for decay ranged from 0 for sound teeth, 1-2 for early non-cavitated lesions, to 3-6 for cavitated lesions. The dental hygienists were calibrated against a gold standard dentist expert. The inter-rater reliability for the hygienists, i.e., weighted kappa, ranged from 0.67 to 0.83 for decay, indicating good to excellent inter-rater reliability and an unweighted kappa of 0.96 to 0.99 for fillings, indicating almost perfect agreement.^28^ In the bivariate analysis, both the count of cavitated decayed primary teeth (ICDAS lesion code 3-6) and a dichotomized variable, i.e. “yes” (>= 1) and “no” (0) were reported. The number of primary teeth of each tooth type with treatment need [anterior (incisors and canines) or posterior (molars)] and number of quadrants (upper right and left and lower right and left) with decayed teeth were also reported.

Surface-level baseline untreated decay on primary teeth (dt) was visualized using odontograms, which display the decay proportion for each of the five surfaces (buccal [B], lingual [L], mesial [M], distal [D], and occlusal [O]) of each primary tooth. Odontograms were constructed separately for the upper and lower arches, with teeth arranged from second right molar (R5) to second left molar (L5).^15^

#### Medicaid Data

Claims data were obtained through a data use agreement between University Hospitals Cleveland Medical Center and the Ohio Department of Medicaid (D-1617-05-0548) and were used to assess the level of preventive and restorative treatment, plus extractions, each child received. Dental Utilization was defined as having any preventive (diagnosis, D0110-D0330; prevention, D1110-D1351; D4355) or restorative dental claim. A full list of assumptions for coding dental attendance has been previously published eTable 2.^23^ A restorative/extraction treatment was identified as the presence of CDT codes D2000-2999 (restorative procedures), which included D2100-2199 (amalgam), D2300-2399 (resin-based composite), and D2930-2933 (crown) or an extraction (D7111-D7210). The treatment codes used to identify the treated teeth for each child are in Supplemental Table 1. Operating room use was identified as the presence of CDT codes D9420, D9219, D9220, D9221, and D9223; or the presence of CPT codes (41899, or 00170) plus a restorative/extraction CDT code. If no OR use codes were present, then it was assumed that treatment occurred in the outpatient setting.

#### Outcomes

Treatment receipt was defined as the difference in the number of teeth treated according to Medicaid Claims minus the number of untreated decayed teeth from the baseline dental exams. Negative values indicated less treatment compared to the number of baseline decay (treatment gap), while positive values and zero were considered adequate treatment receipt.

#### Statistical Analysis

All statistical analyses were conducted using R, version 4.6.0 (R Group for Statistical Computing), RStudio, version 2026.04.0 Build 526. Bivariate analyses (t-test and χ2 test) were used to investigate the differences between those who received adequate treatment and those with treatment gap from the Medicaid data within 1 year.

Multivariable logistic regression was conducted to assess factors associated with the binary outcome of treatment gap (coded 1) versus adequate treatment receipt (coded 0). Covariates included the child’s age, sex, race, caregiver education, and the number of teeth of each tooth type with treatment need (anterior or posterior). For the multivariable analysis, it was assumed that the data were missing at random (MAR), and a complete case analysis was conducted. Odds ratio coefficients and 95% confidence intervals (CIs) were presented for the model.

## Results

Of the 1023 children enrolled in the main clinical trial, 257 were excluded for reasons shown in Figure 1, while 766 had continuous Medicaid enrollment for 1 year and completed the baseline clinical exam. Of those, 487 (63.6%) had a Medicaid claim for dental visit.

**Figure 1:**
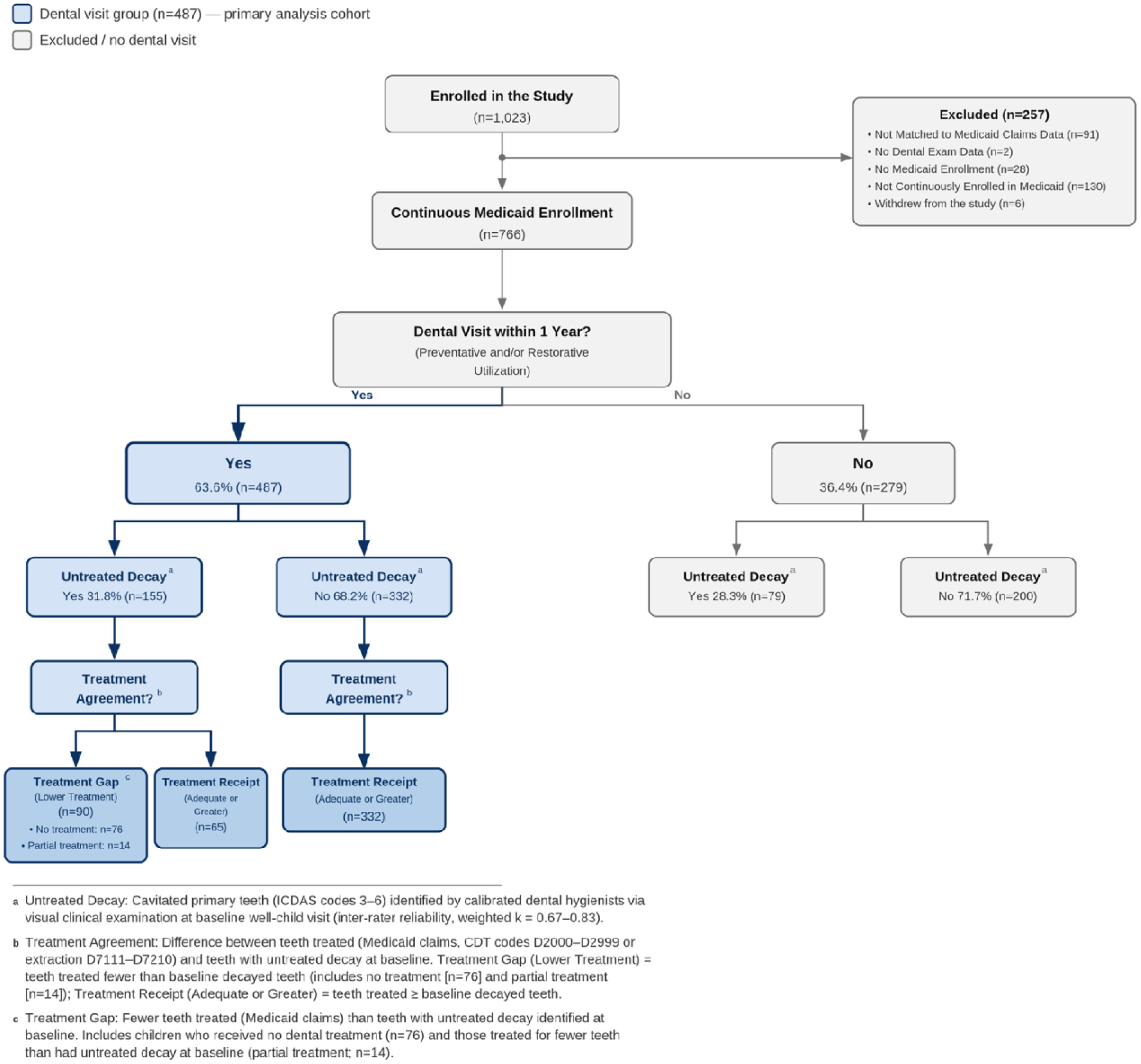
Flowchart of Dental Attendance, Untreated Decay (Hygienists’ Clinical Exams), and Treatment Reciept (Medicaid Claims) in a cohort of 3–6-Year-Old Children Over a 1-Year Period.

Of those, 155/487 (31.8%) had untreated tooth decay, and 65/155 (41.9%) had adequate or greater treatment and 90/155 (58.1%) had a treatment gap with 76/155 (49.0%) getting no treatment at all. (Figure 1 and Table 1)

**Table 1.**
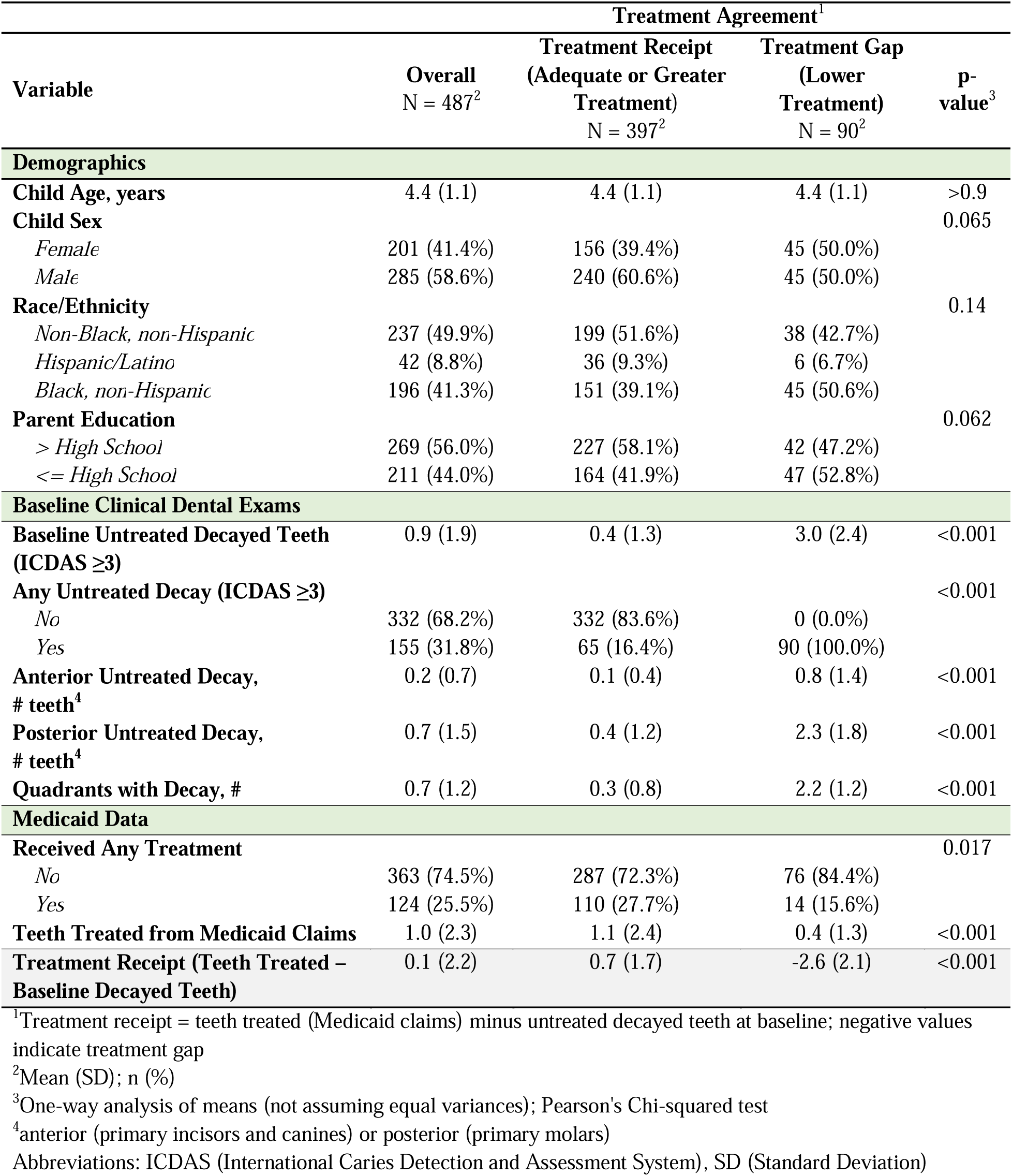
Participant Characteristics Stratified by Treatment Agreement.

For those who received any treatment, the number of teeth treated from Medicaid claims data was average 4.56 ± 3.4. About 63.7% (79/124) received treatment in the outpatient setting, while 36.3% (45/124) received it in the OR. Those who were treated in the OR vs. an Outpatient setting had higher mean primary decayed teeth at baseline (3.6 ± 2.8 vs. 1.0 ± 1.4, p-value = <0.001). The types of treatments differed by setting, with more resin in the outpatient setting, and more stainless-steel crowns in the OR. (Supplemental Table 1)

Overall, the surface level odontograms for baseline untreated decay illustrate that children with a treatment gap carried a higher and more widespread surface-level caries burden, especially in the upper anterior teeth and the occlusal surfaces of the posterior teeth. Among those with a treatment gap, surfaces with the highest proportion of baseline decay were the occlusal surfaces of the primary molars in both arches, which exfoliate around 9–12 years^15^, with decay proportions of about 0.3. The buccal and lingual surfaces of the upper anterior teeth also showed elevated decay in the treatment gap group and these teeth exfoliate earlier, around ages 6–8 years ^15^. (Figure 2)

**Figure 2:**
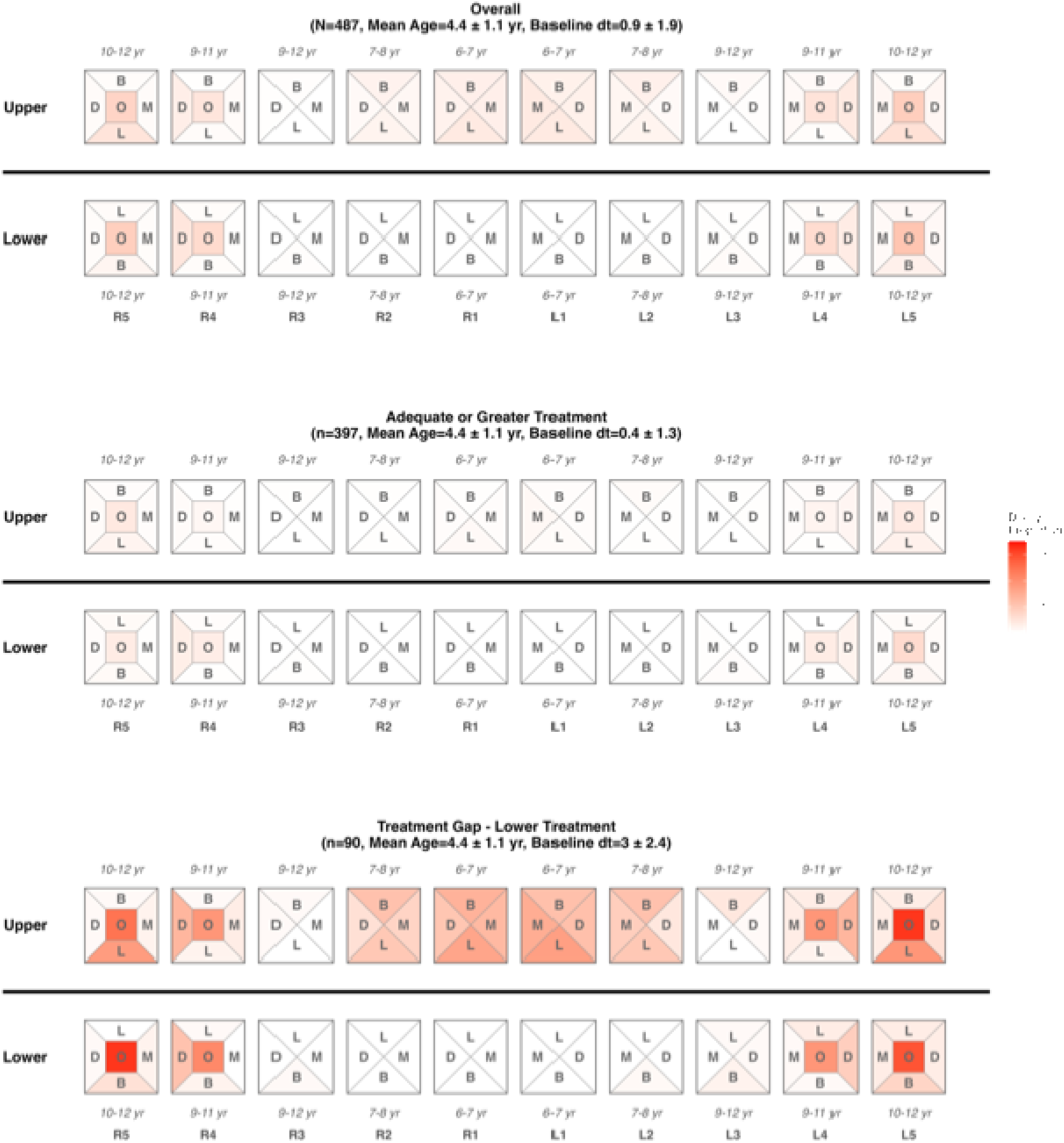
Odontograms of Untreated Decay (International Caries Detection and Assessment System ≥ 3) in Primary Teeth by Surface, Stratified by Treatment Receipt (Treatment Received within 1y Relative to Baseline Untreated Decay), Annotated with Exfoliation Ages of Each Tooth Footnote: Each diagram represents a primary tooth crown viewed from the occlusal perspective, divided into five surfaces. Teeth are arranged by arch position from right second primary molar (R5) to left second primary molar (L5), with upper and lower arches displayed separately. Age ranges above each tooth indicate the typical exfoliation timing for that tooth. Color intensity reflects surface-level untreated decay proportion, with deeper red indicating greater decay proportion. Analyses are presented for three groups: the overall sample (n =487) , children who received adequate or greater dental treatment (n=397), and children classified having a treatment gap (lower treatment) (n=90). All groups had a comparable mean age of 4.4 ± 1.1 years. Those who had a treatment gap had significantly more untreated decay at baseline (International Caries Detection and Assessment System Decay Code ≥ 3) and the decay was present in the highest proportion on the upper anterior teeth and upper and lower molars especially on the occlusal surface. Abbreviations: N = total sample size; n = subgroup sample size; yr = year(s); dt = decayed primary teeth; B = Buccal; D = Distal; O = Occlusal; M = Mesial; L (tooth surface) = Lingual

In the multivariable logistic regression model, number of anterior and posterior decayed teeth at baseline were the strongest predictor of treatment gap. Each additional anterior tooth with baseline untreated decay was associated with 2.19 times the odds of a treatment gap (95% CI: 1.51–3.39; p < 0.001), and each additional posterior decayed tooth was associated with 1.90 times the odds of a treatment gap (95% CI: 1.60–2.30; p < 0.001). Sociodemographic variables were not significantly associated with a treatment gap in this model (Figure 3).

**Figure 3:**
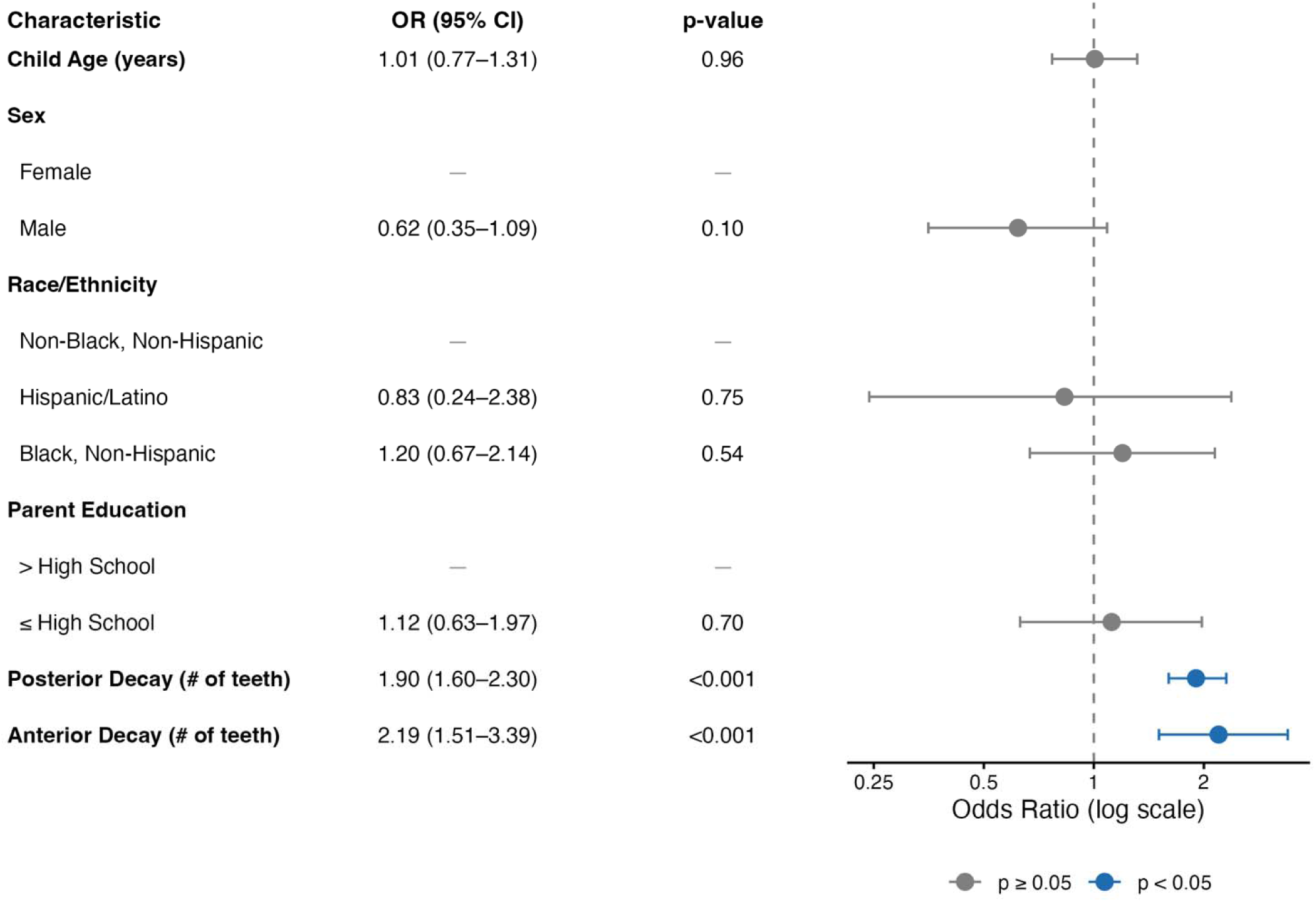
Multivariable Logistic Regression for Treatment Gap (Lower Treatment) adjusted for demographics and number of teeth with decay at baseline (anterior and posterior) (n = 470) Multivariable logistic regression was conducted for Aim 2, with the binary outcome coded as treatment gap (fewer teeth treated than baseline untreated decay; coded 1) versus treatment receipt (adequate or greater treatment received; coded 0). The model covariates included the child’s age, sex, race, caregiver education, and the number of teeth of each tooth type with treatment need [anterior (primary incisors and canines) or posterior (primary molars)] Abbreviation: CI = Confidence Interval, OR = Odds Ratio

## Discussion

In this cohort of 3-6 year old Medicaid enrolled children attending well-child visits, overall 64% had a dental visit based on claims data and this is higher than recent national estimates.^6^ Utilization alone does not fully account for oral health disparities in this population. Even among dental attenders, only 42%│ those with untreated decay from clinical exams received treatments within 1 year that aligned with their baseline dental needs.

A treatment gap may partially reflect the clinical judgement of the outpatient dentist. A dentist might have been hesitant to restore teeth approaching exfoliation, given that the follow-up period in our study intersected with the typical exfoliation age of 6-8 years old for upper anterior teeth.^15^ As a check, we looked at all teeth examined both at baseline and the one-year follow-up dental exam, and only 3.3% had been lost (extraction, or natural exfoliation) indicating minimal impact on our treatment gap estimates. Our finding that having more anterior teeth with untreated decay was associated with a treatment gap is consistent with documented decision-making patterns of pediatric dentists reported. A 2014 survey of 723 pediatric dentists also found that they were most hesitant to restore an anterior primary tooth at or near the age of exfoliation.^29^ To fill this gap, pediatric providers can apply SDF to anterior teeth at well-child visits and counsel caregivers that the resulting dark staining is a visible marker of arrested tooth decay and that SDF lowers cariogenic bacterial load^30^ and may prevent bacteria from attacking the newly erupting permanent teeth. Although primary anterior teeth will exfoliate soon, parents may still prefer a dental referral from primary care providers to cover up SDF staining with a tooth-colored material.^31^

We also observed a significant relationship between a higher number of posterior decayed teeth and higher odds of having a treatment gap. Primary molars do not exfoliate until between 9-12 years of age and thus one would expect to see restoration in 3–6-year-olds who visit the dentist. Child cooperation has been identified as the most important factor influencing dentists’ willingness to restore posterior primary teeth.^32^ In addition, dentist have variability in restorative treatment thresholds for primary molar caries. ^33^ SDF application by pediatric primary care providers may be especially valuable for posterior primary teeth in young children as these teeth have a longer lifespan, and staining is largely hidden on posterior surfaces.

A treatment gap was not significantly related to child’s age, race/ethnicity, or parent’s education but as the number of anterior and posterior teeth with decay increased so did the odds that the child had a treatment gap (Figure 3) and this has implications for caregivers. For children with extensive decay, we speculate that caregivers might find it more challenging to secure necessary follow-up appointments for treatments due to well-known barriers such as the limited number of dentists who accept Medicaid and who practice in low-income neighborhoods^34^ and prolonged wait times for appointments.^35^ Further, the concentration of decay on upper anterior teeth and molar occlusal surfaces is consistent with a tooth decay pattern driven by carbohydrate containing liquids pooling on upper anterior teeth during feeding or “sippy cup” use and lack of appropriate tooth brushing leading to plaque retention in molar pits and fissures.^36,37^ Pediatric primary care providers can counsel caregivers to avoid prolonged exposure to liquids containing sugar including milk, formula and fruit juice, and counsel on daily tooth brushing with a “pea-sized” amount fluoridated tooth paste^37^.

Among young, Medicaid-enrolled children in the United States, treatment of tooth decay is one of the most common indications for elective GA^38^ and pediatricians routinely complete pre-operative clearances for these procedures^39^. Our findings indicate that children treated in OR had more caries. The OR might be a source of delay in care and thus disease progression. This could be due to the fact that children visiting general dentists are not able to get treatment and therefore delay care until they are able to get treatment by scarce pediatric dentists. In fact, a survey of dentists affiliated with Medicaid managed care in New York City found that only 34% were willing to treat 3-5-year-olds with complex restorative needs.^13^ Since caries in young children can rapidly progress if left untreated, ^40^ relying on scarce pediatric dentists who accept Medicaid is not an effective solution. In addition, publicly insured programs such as Medicaid incurred the brunt of the higher costs. A 2018 cost estimate in the US found that the average cost for treating dental decay in young Medicaid-enrolled children was $12,531 for treatment in the OR with GA, and $1,842 for treatment in the outpatient setting.^20^ SDF application by pediatric primary care providers at well-child visits could be an effective intervention to reduce the need for dental restorations in OR with GA as shown in a previous study.^41^

There are limitations to our data: First, children who did not have any baseline decay could have developed new or severe decay within one year at the time of their dental visit. Previous literature has shown that in less than a year a lesion can progress to the point that treatment is required^40^; Second, the treatment gap could have been influenced by parent-related barriers such as lack of transportation and childcare support or delay in seeking care due to the COVID-19 pandemic; Third, the dental hygienists in the study could have over- or under- estimated the presence of cavities as they were relying only on visual methods to detect caries. Although not supported by the literature ^42^, it is possible that without radiographs the hygienists might have missed proximal caries requiring treatment. However, this is unlikely to alter findings relating to treatment gaps.

## Data Availability

Deidentified individual participant data will not be made available.

## Conclusion

In sum, our findings indicate more than half of young Medicaid enrolled children attending well-child visits had a dental treatment gap, and there is a need to adopt non-surgical treatments such as SDF in primary care to address treatment gaps. Sharing responsibility among pediatric providers (pediatricians, pediatric nurse practitioners and nurses), dental hygienist, and dentists, and allowing reimbursement for early interventions like SDF, can reduce this treatment gap and the use of costly OR treatments for Medicaid-enrolled children. In fact, the AMA in 2023 approved a new CPT code for medical providers to apply SDF to arrest tooth decay, ^8^ with guidance published by the AAP in 2024 for pediatric clinicians. ^43^ Medical providers can easily apply SDF to visible decay on the anterior teeth and the occlusal surfaces of the posterior teeth, which were had the highest proportion of decay in our study. SDF can also be used to treat primary teeth that are likely to exfoliate soon so that the bacterial load due to caries ^44^ can be controlled and bacteria have less chances of attacking the permanent teeth. Literature shows that children with caries in the primary teeth are 3 times more likely to have future caries in the permanent teeth.^45^ By expanding access to non-invasive and less-burdensome tooth decay treatments at pediatric primary care visits has the potential to address treatment gaps among low-income preschoolers.

## Clinical Trial registry name, registration number

Providers Against Cavities in Children’s Teeth (PACT), ClinicalTrials.gov ID NCT03385629

## Data Sharing Statement

Deidentified individual participant data will not be made available.

## Abbreviations

AAP: (American Academy of Pediatrics)
AAPD: (American Academy of Pediatric Dentistry)
AMA: (American Medical Association)
CDT: (Code on Dental Procedures and Nomenclature)
CI: (Confidence Interval)
CPT: (Current Procedural Terminology)
cRCT: (cluster Randomized Controlled Trial)
dt: (decayed primary teeth)
EMR: (Electronic Medical Record)
GA: (General Anesthesia)
ICD-10-CM: (International Classification of Diseases, 10th Revision, Clinical Modification)
ICDAS: (International Caries Detection and Assessment System)
MAR: (Missing At Random)
NHANES: (National Health and Nutrition Examination Survey)
NIDCR: (National Institute of Dental and Craniofacial Research)
OR: (Operating Room / Odds Ratio)
PACT: (Providers Against Cavities in Children’s Teeth)
SDF: (Silver Diamine Fluoride)
STROBE: (Strengthening the Reporting of Observational Studies in Epidemiology)
US: (United States)
WCV: (Well-Child Visit)

---

**Supplemental Table 1:**
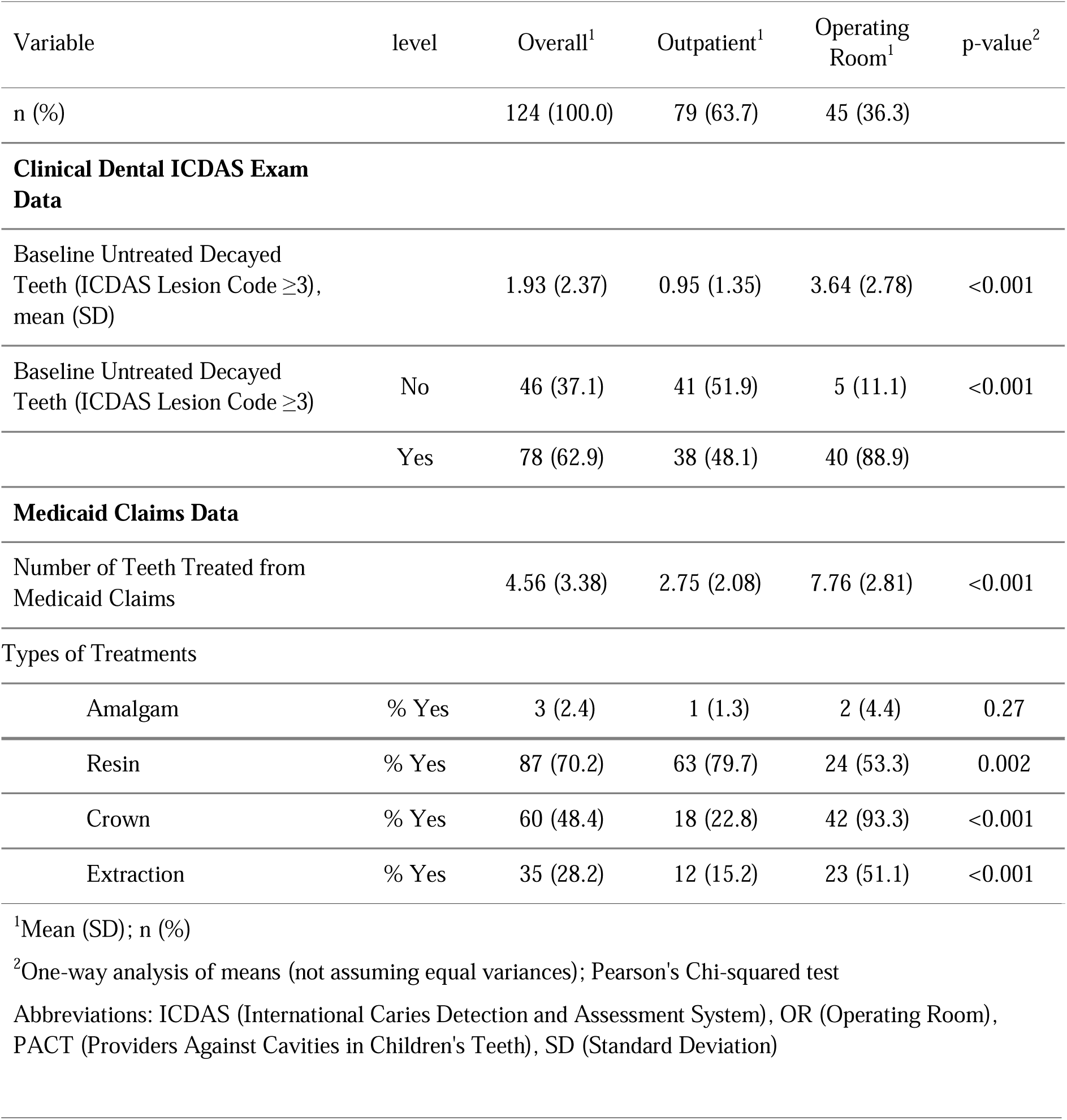
Dental Characteristics of 124 Medicaid Patients Who Received Treatment and Enrolled in the PACT Study Stratified by Treatment in the Operating Room (OR)

