## Supplemental Table 1: for "Treatment Gaps Among Young Medicaid-Enrolled Children with Tooth Decay in Pediatric Primary Care"

| **Supplemental Table 1:** Dental Characteristics of 124 Medicaid Patients Who Received Treatment and Enrolled in the PACT Study Stratified by Treatment in the Operating Room (OR) | | | | | |
| --- | --- | --- | --- | --- | --- |
| Variable | level | Overall^1^ | Outpatient^1^ | Operating Room^1^ | p-value^2^ |
| n (%) |  | 124 (100.0) | 79 (63.7) | 45 (36.3) |  |
| **Clinical Dental ICDAS Exam Data** |  |  |  |  |  |
| Baseline Untreated Decayed Teeth (ICDAS Lesion Code ≥3), mean (SD) |  | 1.93 (2.37) | 0.95 (1.35) | 3.64 (2.78) | <0.001 |
| Baseline Untreated Decayed Teeth (ICDAS Lesion Code ≥3) | No | 46 (37.1) | 41 (51.9) | 5 (11.1) | <0.001 |
|  | Yes | 78 (62.9) | 38 (48.1) | 40 (88.9) |  |
| **Medicaid Claims Data** |  |  |  |  |  |
| Number of Teeth Treated from Medicaid Claims |  | 4.56 (3.38) | 2.75 (2.08) | 7.76 (2.81) | <0.001 |
| Types of Treatments |  |  |  |  |  |
| Amalgam | % Yes | 3 (2.4) | 1 (1.3) | 2 (4.4) | 0.27 |
| Resin | % Yes | 87 (70.2) | 63 (79.7) | 24 (53.3) | 0.002 |
| Crown | % Yes | 60 (48.4) | 18 (22.8) | 42 (93.3) | <0.001 |
| Extraction | % Yes | 35 (28.2) | 12 (15.2) | 23 (51.1) | <0.001 |
| ^1^Mean (SD); n (%)  ^2^One-way analysis of means (not assuming equal variances); Pearson's Chi-squared test  Abbreviations: ICDAS (International Caries Detection and Assessment System), OR (Operating Room), PACT (Providers Against Cavities in Children's Teeth), SD (Standard Deviation) | | | | | |
